# West Nile Virus Neuroinvasive Disease in Patients Treated with anti-CD20 Therapies

**DOI:** 10.1101/2024.12.08.24318681

**Authors:** Rumyar V. Ardakani, Paul D. Crane, Daniel M. Pastula, Lakshmi Chauhan, Elizabeth Matthews, Kelli M. Money, Anna A. Shah, Amanda L. Piquet, Robert H. Gross, Aaron M. Carlson, Kenneth L. Tyler, John R. Corboy, Enrique Alvarez, Andrew B. Wolf

## Abstract

**Background and objectives:** The literature on severe West Nile virus (WNV) neuroinvasive disease (WNND) in patients treated with anti-CD20 therapies is limited to case reports. We systematically characterize cases of WNND in the tertiary academic UCHealth system.

**Methods:** A retrospective cohort (January 2016 to January 2024) of patients with a validated diagnosis of WNND and anti-CD20 medication use was identified with electronic medical record (EMR) query followed by individual chart review.

**Results:** We identified 25 patients, of whom multiple sclerosis was the most common indication for anti-CD20 therapy in 13 patients (52%). 21 patients (84%) presented with meningoencephalitis. Cerebrospinal fluid (CSF) WNV IgM was positive in 5/21 patients (24%) who were tested, while 13/14 tested patients (93%) had positive RT-PCR findings in the CSF. MRI demonstrated anomalies associated with WNND in 12/23 patients (52%) with available imaging. ICU admission was required in 8 patients (32%) and 12 patients (48%) were treated with intravenous immunoglobulin. Worsening of ≥1 point from pre-WNV baseline modified Rankin scale (mRS) to the 90-day post-discharge mRS was seen in 18 patients (75%). Two patients (8%) died by 90-days.

**Discussion:** WNND leads to disability accrual in patients on B-cell depleting anti-CD20 therapies. Utilization of RT-PCR is important in optimizing diagnosis due to limited sensitivity of antibody testing.

## INTRODUCTION

West Nile virus (WNV) is a mosquito-borne flavivirus that causes a range of clinical manifestations.^1^ While many WNV infections are asymptomatic or result in a relatively mild febrile illness, rare neuroinvasive cases resulting in encephalitis, meningitis, and/or myelitis result in substantial morbidity and mortality.^2–4^ Between 1999 and 2017, over 22,000 WNV neuroinvasive disease (WNND) cases were reported in the United States with a mortality rate estimated at 10%.^5^ Immunosuppressed patients, including those with iatrogenic immunosuppression, are at elevated risk of developing severe WNV infection and having poor outcomes should they become infected.^6,7^

Anti-CD20 monoclonal antibodies have been increasingly used for oncological, rheumatological, and neuroimmunological indications. Anti-CD20 therapies may increase risk of infections via B-cell depletion and potential development of hypogammaglobulinemia with sustained use.^8^. A study in the Mayo Clinic system has identified that patients who are immunosuppressed across a broad range of indications have higher rates of mortality and ICU admission from WNND than immunocompetent patients.^7^ Patients on anti-CD20 therapies for any indication who develop arboviral infections (predominantly WNV) have fatality rates reported as high as 79% based on recent literature review.^6^ In part, the severity of WNV infection in these patients may be driven by the importance of humoral immune response in combating the infection.^6,9–11^ However, there may be bias impacting such case studies as patients with milder illness may not be reported. WNV diagnosis may also prove more challenging in patients on anti-CD20 therapies due to potential false-negative antibody testing in the setting of B-cell depletion impairing serological response.^12,13^

Despite the severity of WNV infections in anti-CD20 treated patients, risk factors are not robustly described.^1,14^ There is limited data on clinical outcomes of WNND in anti-CD20 treated patients and no targeted therapies are available. Better understanding of this population is important to help develop improved diagnostic and treatment strategies. Here we evaluate outcomes of WNND in a retrospective cohort of patients treated with anti-CD20 therapies.

## METHODS

### Regulatory Approvals

This retrospective cohort study was reviewed by the Colorado Multiple Institutional Review Board (COMIRB) and exempted based on secondary use of deidentified data. Patient consent was not required.

### Patient Identification

Patients were identified in the UCHealth Epic electronic medical record (EMR) using the Slicer-Dicer tool. UCHealth is a multisite healthcare system operating in Colorado, Wyoming, and Nebraska with 14 acute-care hospitals and several dozen clinic locations. EMR searches were completed for a period spanning January 1, 2016, to January 1, 2024. Searches were completed for patients with ICD-10 diagnostic codes for WNV infection (A92.30, A92.31, A92.30) or any abnormal serum WNV immunoglobulin (Ig) M (IgM), cerebrospinal fluid (CSF) WNV IgM, serum WNV reverse transcription polymerase chain reaction (RT-PCR), or CSF WNV RT-PCR with documented use of anti-CD20 therapy including rituximab (Rituxan and biosimilar products Riabni, Ruxience, and Truxima), ocrelizumab (Ocrevus), ofatumumab (Arzerra, Kesimpta), ublituximab (Briumvi), and obinutuzumab (Gazyva).

### Inclusion and Exclusion Criteria

All identified charts were reviewed by a board-certified neurologist (RVA, PDC, or ABW) to confirm diagnosis of WNND. Patients aged 18-85-years were included. Diagnosis required the presence of a typical clinical syndrome, laboratory evidence of infection, and exclusion of alternative causes at the discretion of the chart reviewer. Typical clinical syndromes were defined as meningitis (headache, fever, nuchal rigidity), encephalitis (altered mental status, seizures, movement disorders), myelitis (motor or sensory deficits localized to spinal cord), or some combination of these syndromes, notably meningoencephalitis. Laboratory evidence of infection required positivity of ≥ 1 of serum WNV IgM, CSF WNV IgM, serum WNV RT-PCR, or CSF WNV RT-PCR. Patients with absent or negative laboratory values but with WNND diagnosis per diagnosis codes or clinical notes were excluded. Charts were also reviewed for details on the use of anti-CD20 therapies including timing, dosing, and laboratory monitoring.

Given standard dosing intervals and the typical duration of efficacy for anti-CD20 therapies, patients were included if there was any dose of anti-CD20 therapy received within 12 months preceding an identified WNV infection or if there was prior anti-CD20 therapy administration with persistent B-cell depletion (<1% of lymphocytes) or absolute lymphopenia (below the laboratory-specific reference range). Patients with anti-CD20 therapy received outside of these parameters were excluded. Inclusion and exclusion criteria were applied to patients identified in the EMR search as in Figure 1.

**Figure 1.**
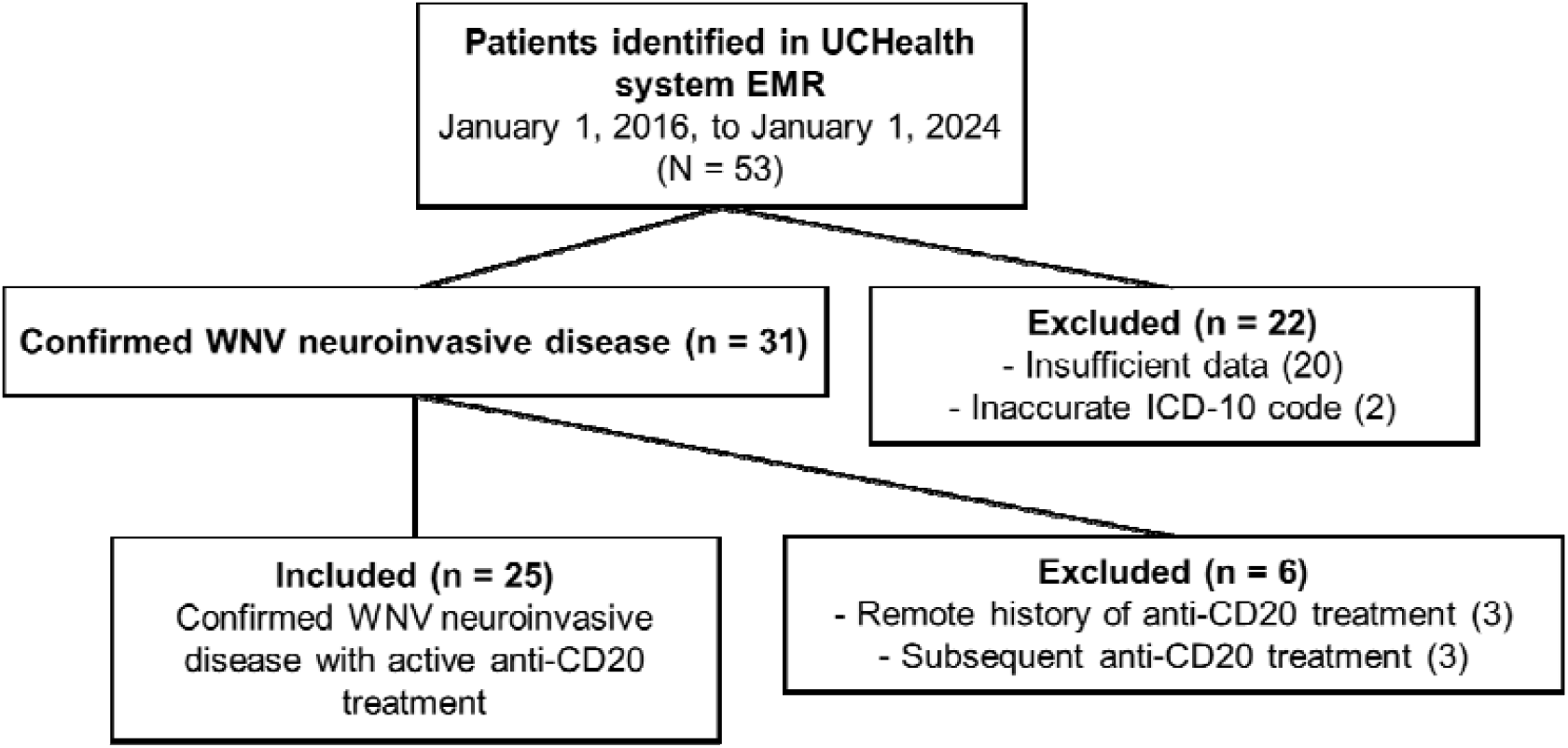
Flow diagram outlining patient exclusion and inclusion based on Electronic Medical Record (EMR) search. WNV = West Nile virus.

### Data Collection

Magnetic resonance imaging (MRI) images and/or reports were classified by the chart reviewers using a structured instrument. Clinical and laboratory data encompassing demographics, clinical signs and symptoms, laboratory values, anti-CD20 use, acute treatment, and clinical outcome were captured in a standardized form. Using a standardized algorithm, the modified Rankin score (mRS) was retrospectively captured for each patient at timepoints including pre-WNV baseline, at time of WNV symptomatic presentation, at time of hospital discharge, and at the follow-up visit closest to 90-days from discharge.^15^

### Outcome Measures

Development of severe WNND (defined as a composite of death or ICU admission) was the primary outcome with exploration of potential risk factors age, IgG levels, and lymphopenia. Patients were stratified by age ≥ 50-years-old vs age < 50-years-old based on prior evidence demonstrating elevated mortality from WNV in patients ≥ 50-years-old; by baseline IgG < 500 mg/dL vs. baseline IgG ≥ 500 mg/dL due to previously demonstrated increase risk for infections with this degree of hypogammaglobulinemia when on rituximab; by baseline disability mRS 0-1 (no functional impairment) vs MRS ≥ 2 (functional impairment); and by preceding lymphopenia (below laboratory reference range).^16,17^ Clinical improvement was defined as mRS improvement ≥1 point from admission to 90-days post-discharge while clinical stability or worsening was defined by mRS with no change or worsening ≥1 point from admission to 90-days post-discharge.

### Statistical Analysis

Statistical analyses were completed in Microsoft Excel (Redmond, WA) and IBM SPSS (Chicago, IL). Clinical and demographic features were reported as frequencies for categorical variables and mean or median with standard deviation or interquartile range for continuous variables. Fisher’s exact test was used for comparison of clinical outcome measures with two-tailed significance defined at 0.05.

### Data Sharing

Anonymized data not published within this article will be made available by request from any qualified investigator with a data-sharing agreement.

## RESULTS

### Demographics & Baseline Characteristics

53 patients were identified in the EMR search, of which 28 were excluded, primarily due to insufficient laboratory data or clinical documentation to confirm diagnosis (20 patients), while 6 patients received anti-CD20 treatments outside of the temporal requirements, and 2 patients had inaccurate ICD codes [Figure 1]. 25 patients were included with mean age of 54.3 years (standard deviation 12.4 years) and median mRS 1 (interquartile range [IQR] 1-2) [Table 1].

**Table 1.** Demographics and Baseline Characteristics. SD = standard deviation, IQR = interquartile range, ALC = absolute lymphocyte count, GPA = granulomatosis with polyangiitis, NMDA = N-methyl-D-aspartate, mRS = modified Rankin scale, IgG = immunoglobulin G

| **Characteristics** | **Patients, Number (%)** |
| --- | --- |
| Age in years, mean (SD) | 54.3 (12.4) |
| Age, median (IQR) | 54 (48-61) |
| ≥ 50 years | 17 (68) |
| < 50 years | 8 (32) |
| Gender |  |
| Male | 10/25 (40) |
| Female | 15/25 (60) |
| Indication for anti-CD20 therapy |  |
| Multiple sclerosis | 13/25 (52) |
| Lymphoma | 5/25 (20) |
| GPA | 4/25 (16) |
| Rheumatoid arthritis | 1/25 (4) |
| NMDA receptor encephalitis | 1/25 (4) |
| Liver transplantation | 1/25 (4) |
| Pre-morbid mRS, median (IQR) | 1 (1-2) |
| mRS 0-1 | 19/25 (76) |
| mRS ≥ 2 | 6/25 (24) |
| Anti-CD20 Therapy |  |
| Rituximab | 13/25 (52) |
| Ocrelizumab | 8/25 (32) |
| Obinutuzumab | 3/25 (12) |
| Ublituximab | 1/25 (4) |
| Anti-CD20 Therapy Duration, median, months (IQR) | 36 (18-70) |
| Serum ALC, median (IQR), 10<sup>9</sup> cells/L | 1.1 (0.9-1.4) |
| Serum CD19+ B cell percentage |  |
| ≤ 1% | 14/17 (82) |
| > 1% | 3/17 (18) |
| Serum IgG level, median (IQR), mg/dL [number missing] | 629 (399-908) [6] |
| IgG ≥ 500 | 12/19 (63) |
| IgG <500 | 7/19 (37) |

Most patients, 13/25 (52%), received anti-CD20 therapy for multiple sclerosis (MS), while 12/25 (48%) received treatment for other indications [Table 1]. Rituximab was used in 13/25 patients (52%), ocrelizumab in 8/25 patients (32%), obinutuzumab in 3/25 patients (12%), and ublituximab in 1/25 patients (4%) with duration of anti-CD20 therapy of 36 months with an IQR of 18-70 months. Serum Immunoglobulin G levels were measured in 19/25 patients preceding or at time of presentation with WNND, with a median of 629 mg/dL (IQR of 399-908 mg/dL) [Table 1]. The median absolute lymphocyte count (ALC) on monitoring blood draws preceding infection was 1.1 (IQR 0.9-1.4 10^9^cells/L).

### Symptomatic Presentation

The most common clinical presentation was meningoencephalitis, which was seen in 24/25 patients (96%) [Table 2]. The most common symptoms included malaise (25/25 patients, 100%), headache (24/25 patients, 96%), and encephalopathy (18/25 patients, 72%), with a range of other symptoms seen in fewer patients [Table 2]. Median mRS at presentation was 4 (IQR 3-4).

**Table 2.** Clinical presentations & paraclinical data for anti-CD20-treated patients with WNV neuroinvasive disease. CSF = cerebrospinal fluid, IQR = interquartile range, mRS = modified Rankin scale, WNV = West Nile virus, WBC = white blood cell, RT-PCR = reverse transcription polymerase chain reaction, IgG = immunoglobulin G, IgM = immunoglobulin M

| Characteristic | Patients, Number (%) |
| --- | --- |
| Core clinical presentations |  |
| Meningoencephalitis | 21/25 (84%) |
| Meningitis | 24/25 (96) |
| Encephalitis | 21/25 (84) |
| Myelitis | 1/25 (4) |
| Signs and symptoms |  |
| Malaise | 25/25 (100) |
| Nausea, vomiting, and/or diarrhea | 16/25 (64) |
| Fever | 25/25 (100) |
| Headache | 24/25 (96) |
| Encephalopathy | 18/25 (72) |
| Myalgias | 18/25 (72) |
| Cranial neuropathy | 3/25 (12) |
| Rash | 4/25 (16) |
| Tremor | 3/25 (12) |
| Seizure | 6/25 (24) |
| Weakness | 14/25 (56) |
| Radiculitis | 1/25 (4) |
| Vestibular neuritis | 1/25 (4) |
| Imaging abnormalities |  |
| MRI brain abnormalities | 12/25 (48) |
| MRI spine abnormalities | 1/10 (10) |
| Diagnostic labs |  |
| Serum WNV IgM positive | 7/16 (44) |
| Serum WNV IgG positive | 1/16 (6) |
| Serum WNV RT-PCR positive | 8/8 (100) |
| CSF WNV IgM positive | 5/21 (23) |
| CSF WNV IgG positive | 1/21 (5) |
| CSF RT-PCR positive | 13/14 (93) |
| CSF WBC, median (IQR) [number missing] cells/mm<sup>3</sup> | 33 (20-128) [4] |
| CSF protein, median (IQR) [number missing] mg/dL | 87 (67-110) [4] |
| CSF glucose, median (IQR) [number missing], mg/dL | 56 (51-68) [4] |
| mRS at presentation, median (IQR) | 4 (3-4) |

### Laboratory Testing

Serum WNV IgM was positive in 7/16 tested patients (44%) while CSF WNV IgM was positive in 5/21 patients (24%) [Table 2]. Serum WNV RT-PCR was positive in 8/8 tested patients (100%), while 13/14 tested patients (93%) had positive RT-PCR findings in the CSF [Table 2]. CSF analysis revealed a median white blood cell count of 33 cells/µL (IQR 20-128 cells/µL), median protein 87 mg/dL (IQR 67-110 mg/dL), and median glucose 56 mg/dL (IQR 48-68 mg/dL) [Table 2]. CSF had a lymphocytic pleocytosis (>70% of nucleated cells) in 9/20 patients, neutrophilic pleocytosis (>70% of nucleated cells) in 1/20 patients, and mixed findings in 10/20 patients.

### Imaging

MRI demonstrated anomalies associated with WNND in 12/23 patients (52%) with brain imaging and in 1/10 patients (10%) with spinal cord imaging [Table 3, Figure 2]. The majority of findings were new T2 FLAIR hyperintensity (12/23, 52%), with 9/23 (39%) demonstrating gadolinium enhancement and 2/23 (9%) demonstrating diffusion restriction [Table 3].

**Table 3.** MRI patterns of WNV neuroinvasive disease in anti-CD20-treated patients. Multiple anatomical regions may

| **Finding** | **Patients, Number (%)** |
| --- | --- |
| Acute T2 FLAIR hyperintensity | 12/23 (52) |
| Brainstem | 1/23 (4) |
| Thalamus | 5/23 (22) |
| Temporal lobe | 1/23 (4) |
| Basal ganglia | 2/23 (9) |
| Cortex | 4/23 (17) |
| Cerebellum | 5/23 (22) |
| Spinal cord | 1/10 (10) |
| Contrast enhancement | 9/23 (39) |
| Brainstem | 1/23 (4) |
| Thalamus | 2/23 (9) |
| Temporal lobe | 0/23 (0) |
| Basal ganglia | 1/23 (4) |
| Cortex | 1/23 (4) |
| Cerebellum | 3/23 (13) |
| Spinal cord | 1/10 (10) |
| Meningeal | 8/23 (35) |
| Nerve root | 2/23 (9) |
| Diffusion restriction | 2/23 (9) |
| Brainstem | 0/23 (0) |
| Thalamus | 1/23 (4) |
| Temporal lobe | 0/23 (0) |
| Basal ganglia | 1/23 (4) |
| Cortex | 0/23 (0) |
| Cerebellum | 1/23 (4) |

**Figure 2.**
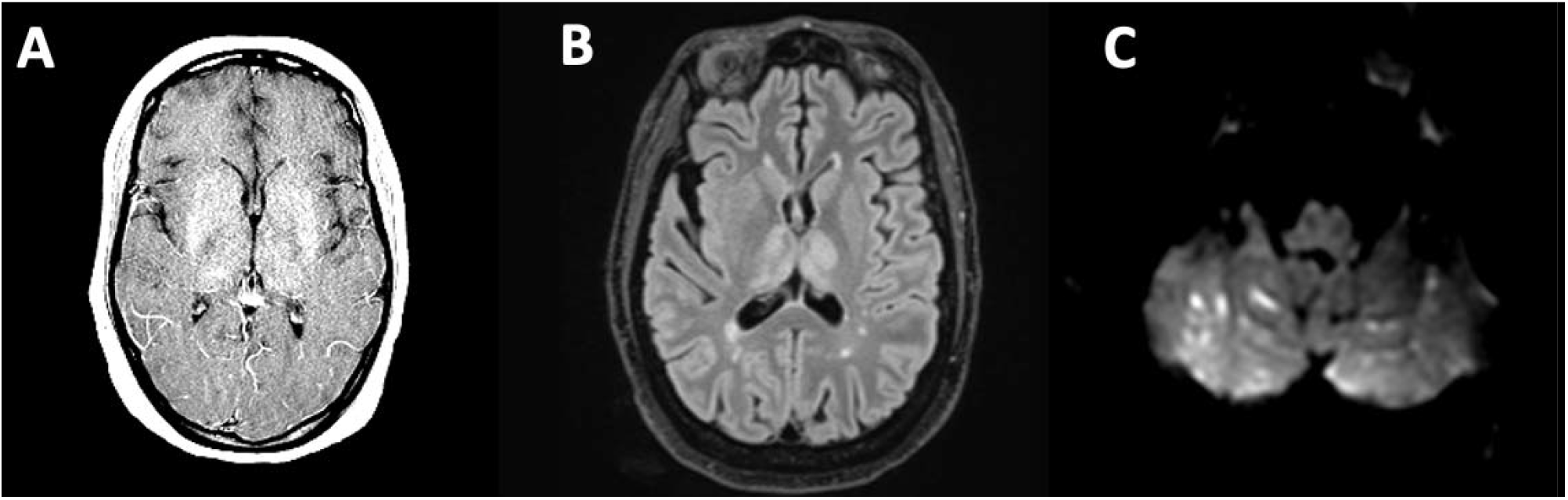
Selected images of characteristic MRI patterns of anti-CD20-treated patients with West Nile virus neuroinvasive disease. A.) T1 post-gadolinium sequence demonstrating symmetrical diffuse contrast enhancement of the bilateral basal ganglia and thalamus; B.) T2 FLAIR sequence demonstrating symmetrical hyperintensities involving the bilateral thalami on background of chronic demyelinating lesions; C.) Diffusion weighted sequencing sequence demonstrating right greater than left cerebellar diffusion restricting lesions (apparent diffusion coefficient sequence with corresponding hypointensity not shown).

### Treatment & Clinical Outcomes

IVIg treatment was given in 12/25 patients (48%) [Table 4]. ICU admission was required in 8/25 patients (32%), with 6/25 patients (24%) requiring mechanical ventilation [Table 4] The median hospital stay was 13 days (IQR 5-37 days) [Table 4]. The median discharge mRS was 3 (IQR 3-4). The median estimated 90-day post-discharge mRS was 3 (IQR 2-4) with 24/25 patients (96%) assessed at median date of 91.5 days post-discharge (IQR 80-108 days).

**Table 4.**
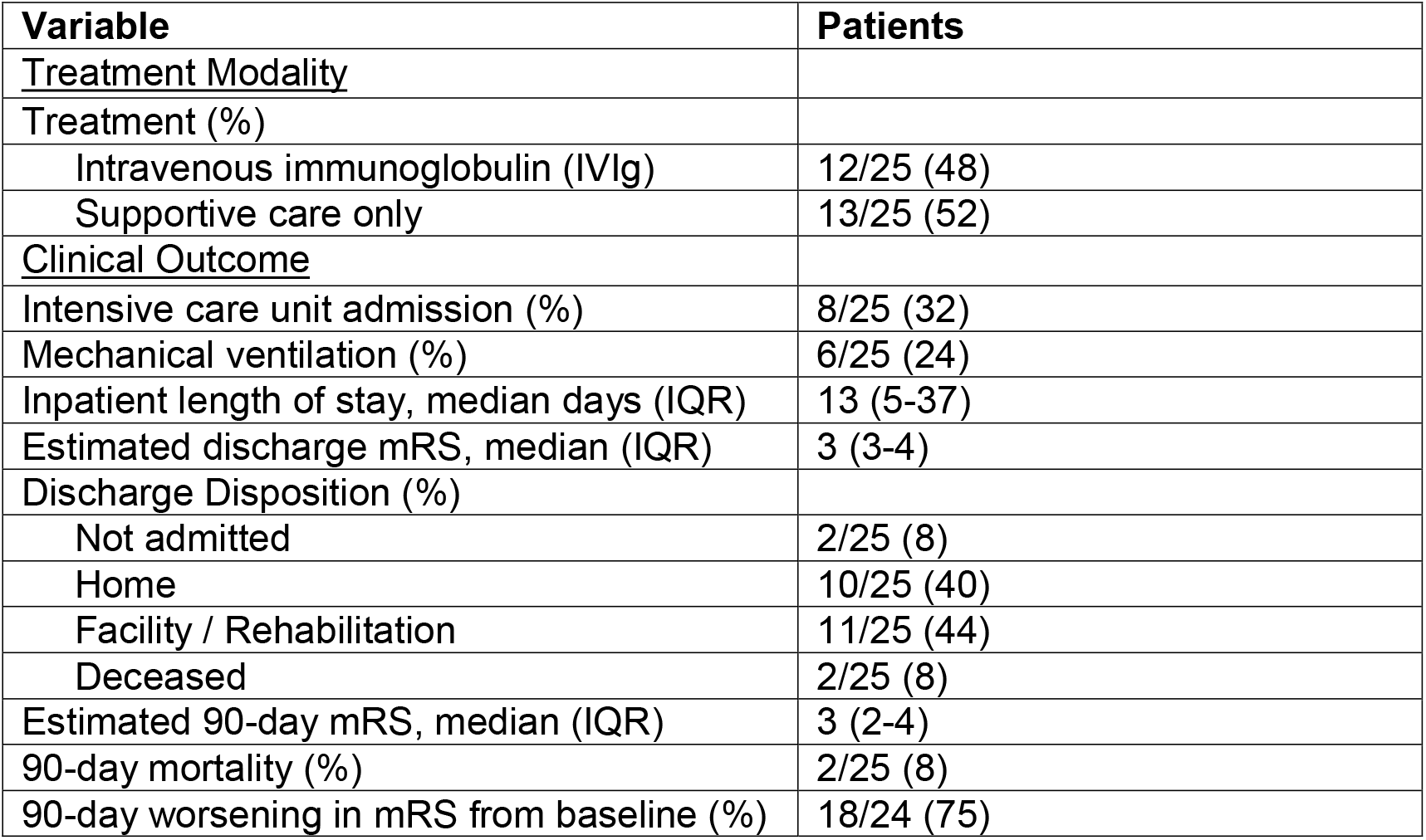
Treatment Modalities and Clinical Outcomes for WNV neuroinvasive disease. IQR = interquartile range, mRS = modified Rankin Score, IVIg = intravenous immunoglobulin

Worsening of ≥1 point from pre-WNV baseline mRS to the 90-day post-discharge mRS was seen in 18/24 patients (75%). Compared to the admission mRS, 15/24 patients (62%) improved ≥1 point on 90-day post-discharge mRS, while 7/24 patients (29%) were stable, and 2/24 patients (8%) worsened, both of whom died. The 90-day mortality was 2/25 (8.0%) [Table 4].

Two patients died from WNV, one was being treated for MS and the other for follicular lymphoma. The patient treated for follicular lymphoma was receiving treatment with obinutuzumab monotherapy and had profound hypogammaglobulinemia (IgG 223mg/dL) and persistent lymphopenia (ALC 0.2 × 10^9^ cells/L) dating to at least 6 months prior to infection with accompanying CD19+ depletion. The patient with MS was being treated with rituximab monotherapy and had an IgG of 939 but persistent lymphopenia (ALC 0.4 × 10^9^ cells/L) dating to at least 6 months prior to infection with accompanying CD19+ depletion.

Of the 25 identified patients, risk factors for severe WNND were evaluated. There was no association of age ≥50-years-old (p=1.000), IgG <500mg/dL (p=0.320), pre-existing lymphopenia (p=0.081), or baseline MRS ≥ 2 (p=0.059) with developing severe WNND. Sample sizes precluded multivariate analyses.

Patients treated with IVIg did not differ from those who were not treated with IVIg by age (p=0.202), IgG <500mg/dL (p=1.000), or pre-existing lymphopenia (p=.668). IVIg treatment was not associated with mRS improvement ≥1 point from admission to 90-days post-discharge (p=0.415).

## DISCUSSION

In an area of limited data, we demonstrate the substantial impact of WNND on disability accrual for patients immunosuppressed with B-cell depleting anti-CD20 therapies and support the need for RT-PCR testing for accurate diagnosis in this population. We build on previous studies by systematic detection of cases from an integrated health system over an extended time-period.

CSF WNV RT-PCR was more sensitive than serological testing (WNV IgM) in establishing the diagnosis of WNND in these patients on anti-CD20 therapies. This has previously been described in case series and is now supported by this systemic study.^13^ In fact, most patients in this cohort had negative WNV serological testing and thus the diagnosis was only able to be established with RT-PCR. This is contrary to patients who are not on anti-CD20 therapy in whom serological testing is typically considered to be more sensitive than RT-PCR due to short-lived viremia in WNV infection.^12^ This may reflect an impaired ability for patients on anti-CD20 therapies to mount a robust humoral response due to B-cell depletion. Therefore, when WNND is suspected in patients on anti-CD20 therapy, it is critical to use RT-PCR for evaluation.

The degree of CSF pleocytosis was generally less than what is reported in patients with WNND (mean 33 cells/uL in this study vs 226 cells/uL in a previous study of 250 patients), which may reflect impaired immune response in patients who are on anti-CD20 therapy.^18^ Consistent with prior findings, the most common regions of involvement demonstrated on MRI included the deep gray nuclei (thalamus and basal ganglia) and cerebellum.^19^ However, a sizable number (48%) of patients did not have any changes identified on MRI, demonstrating that imaging can have limited sensitivity, although this may be impacted by timing of MRI from symptom onset.

We show that patients who develop WNND on anti-CD20 therapies are frequently critically ill in the acute setting and can accumulate significant disability that persists post-discharge. As such, improved treatments of WNND and methods to reduce risk are important. There are currently no approved treatments for WNND and supportive care remains the mainstay of treatment. The largest clinical trial assessing the efficacy of IVIg and Omr-IgG-am (immunoglobulin product with high titers of anti-WNV antibodies) did not demonstrate benefit.^14^ However, this study was designed to evaluate safety and was underpowered for detection of morbidity and mortality outcomes.^14^ This study did not differentiate between immunocompromised and immunocompetent patients. Immunocompromised patients are less likely to mount a humoral response to WNV infection and thus may potentially benefit more from passive transfer of anti-WNV IgG. As such, IVIg is occasionally utilized based on smaller studies suggesting benefit in immunocompromised patients.^20–22^ In the current study, approximately half of patients received IVIg as part of their treatment course. The small sample sizes and heterogeneous population of patients with differing levels of immunosuppression and underlying indications for treatment precluded robust analysis of the impact of IVIg on clinical outcomes

WNV is transmitted by mosquitoes and infection risk may be reduced by limiting exposures. High-risk patients, including those on anti-CD20 therapies, should be counseled on the use of insect repellent, covering exposed skin when outdoors, and limiting time outdoors between dusk and dawn per Centers for Disease Control and Prevention guidance.^23^ Due to the seasonal variation in WNV risk, patients should exercise caution during periods of heightened risks (typically June-November in the United States).

Strengths of this study include the systematic focus on a cohort of patients treated with anti-CD20 therapies, including those with multiple medical conditions, from a state with a high incidence of WNND.^24^ The EMR also allowed for longitudinal capture of laboratory, radiologic, and clinical data. Limitations of the study include its retrospective design making it prone to selection bias and incomplete data availability, especially for patients who were transferred from other institutions or received anti-CD20 therapy outside of UCHealth. While the mRS was designed for stroke clinical trials, it has been used for retrospective studies of WNND and other neurological conditions.^25–27^ However, the mRS may underestimate important deficits and not reflect the dynamic range of disability seen in patients.^25^ It is possible that there was inadequate capture of patients who had WNND due to reliance on ICD-10 codes and laboratory values that are searchable in the EMR. We did not evaluate a comparison group of WNND patients who were untreated or on other immunosuppressive therapies to assess comparative outcomes. A systematic evaluation to better understand the relative risk for developing WNND in patients on different classes of immunosuppression would be a worthy future aim.

This study demonstrates that there is significant morbidity and mortality in patients who develop WNND while on anti-CD20 therapy. The importance of RT-PCR for establishing the diagnosis of WNV in patients on anti-CD20 therapy is also recognized. Given the use of anti-CD20 therapies for a wide array of inflammatory diseases and malignancy, it is critical for clinicians to be aware of unique challenges in the care of patients who develop neuroinvasive WNV infection. This includes the need for the development of targeted treatment strategies for WNV and counseling on preventive measures.

## Notes

### Competing Interest Statement

A.M. Carlson reports grant funding by Amgen which overlapped with the period in which this study was completed.

### Funding Statement

This study did not receive any funding.

